# Genomic epidemiology uncovers the origin of the mpox epidemic in Sierra Leone

**DOI:** 10.1101/2025.10.15.25337823

**Authors:** Allan K.O. Campbell, John Demby Sandi, Ifeanyi F. Omah, Martin Faye, Edyth Parker, Taylor Brock-Fisher, Crystal M. Gigante, Vidalyn Folorunso, Mohamed Saio Kamara, Anu Jegede Williams, Mouhamed Kane, Tiangay Mariama Patience Sallay Kallon, Julian Campbell, Kadiatu Salmata Sesay, Sia Mani, Choe Miller, Naomi Daniel Sesay, Francis Baimba, Mignane Ndiaye, Roberta Lansana, Ibrahim Umaru Fofanah, Simon Ruhweza, Zein Souma, Amanda Kargbo, Komba Koninga, Alie Tia, Jone Ngobeh, Foday Thoronka, Alusine Fofanah, Al Ozonoff, Colby Wilkason, Danny Park, Christopher Tomkins-Tinch, Marietou F Paye, Christopher Shin, Ian Baudi, Brendan Blumenstiel, Patrick Varilly, Ivan Specht, Ben Fry, Karlie Zhao, Paul Cronan, Ellory Laning, Oludayo Oluwaseyi Ope-ewe, Ayotunde Elijah Sijuwola, Femi Saibu, Harouna Soumare, Ebenezer Kehinde Ogundana, Jolly Amoche Adole, Imonikhe Kennedy Kio, Folefac Agnes Njeandoh, Alexandra Tuttle, Walter O. Oguta, Jonathan Greene, Aminata Koroma, Joseph Kanu, Mamadou Aliou Barry, Aboubacry Gaye, Andy Mahine Diouf, Christine Hughes, Joshua I. Levy, Alhaji N’jai, Moussa Moise Diagne, Dolo Nosamiefan, George Ameh, John Klena, Monique A. Foster, Abebaw Kebede, Collins Tanui, Boubacar Diallo, Anise Happi, Sofonias Tessema, Abdourahmane Sow, Yenew Kebede, Isatta Wurie, James Squire, Doris Harding, Zikan Koroma, Mohamed Boie Jalloh, Amadou Alpha Sall, Kristian G. Andersen, Andrew Rambaut, Mohamed Alex Vandi, Ibrahima Socé Fall, Pardis Sabeti, Christian Happi, Foday Sahr, Donald S. Grant

**Affiliations:** National Public Health Agency; Central Public Health Reference Laboratory; Kenema Government Hospital, Sierra Leone; Faculty of Medical Laboratory Sciences and Diagnostics, College of Medicine and Allied Health Sciences, Sierra Leone; School of Community Health Sciences, College of Medical Sciences, Njala University, Sierra Leone; Africa Centres for Disease Control and Prevention (ACDC); Institute of Genomics and Global Health, Redeemer’s University, Ede, Osun State, Nigeria; The Broad Institute of MIT and Harvard, Cambridge, MA, USA; Institute Pasteur de Dakar; Department of Translational Medicine, The Scripps Research Institute, La Jolla, CA, 92037, USA; Fathom Information Design, Boston, MA, USA; Centres for Disease Control and Prevention (CDC), Atlanta, GA, USA; World Health Organisation (WHO), Sierra Leone; Dept. Of Biological Sciences, Fourah Bay College and Dept. Of Microbiology, College of Medicine and Allied Health Sciences, University of Sierra Leone, Freetown, Sierra Leone; Institute of Ecology and Evolution, University of Edinburgh, Edinburgh, UK; Sierra Leone-China P3 Friendship Laboratory; Koinadugu College, Kabala, Sierra Leone; Department of Medical Education, California University of Science and Medicine, Colton, CA; School of Public Health, Njala University, Bo; College of Medicine and Allied Health Sciences, University of Sierra Leone; World Health Organisation Regional Office for Africa (WHO AFRO), Regional Emergency Hub, Dakar - Senegal; Institute for Computational and Mathematical Engineering, School of Engineering, Stanford University, Stanford, CA 94305, USA; Department of Biological Sciences, Faculty of Natural Sciences, Redeemer’s University, Ede, Osun State, Nigeria; Department of Immunology and Infectious Diseases, Harvard T.H. Chan School of Public Health, Harvard University, Boston, MA, USA; Department of Parasitology and Entomology, Nnamdi Azikiwe University, Awka, Anambra State, Nigeria; Department of Clinical Sciences, Liverpool School of Tropical Medicine, Liverpool, UK

**Keywords:** Mpox virus, Clade IIb lineage G.1, Viral transmission dynamics, Sierra Leone outbreak 2025, Bayesian phylogeography, Cryptic circulation, Genomic epidemiology

## Abstract

In January 2025, Sierra Leone reported its first mpox case in eight years, followed by a rapid nationwide surge that made it the epicenter of the continental mpox virus (MPXV) Clade IIb A.2.2 outbreak, with more than 5,000 confirmed cases reported by August. To investigate the origin, timing, and spread of this epidemic, we generated 338 high-quality MPXV genomes from cases across 14 districts and conducted Bayesian phylogeographic analyses. We found that transmission was driven by a newly emerging Clade IIb lineage, G.1, descended from lineages circulating in the ongoing Nigerian epidemic. Phylogeographic reconstructions indicate that G.1 arose in late September 2024 and circulated undetected for approximately three months before establishing sustained transmission in the densely populated Western Area Urban and Rural districts, which became the principal source of repeated viral introductions that drove epidemics in other regions. Together, these findings reveal the origins and dispersal dynamics of the 2025 mpox outbreak in Sierra Leone and underscore that effective control remains achievable through targeted vaccination, strengthened early warning systems, and improved access to genomic and diagnostic surveillance.

## Background

Mpox is a re-emerging zoonotic disease caused by MPXV, which has driven outbreaks across multiple countries in recent years.^1,2^ MPXV is classified into two main clades: I and II. Historically, Clade I was confined to zoonotic infections arising from Central African animal reservoirs, whereas Clade II (subdivided into IIa and IIb) was restricted to West Africa and associated with lower case fatality rates.^3^ In the past decade, MPXV has shifted in both epidemiology and evolution, with large outbreaks in endemic and non-endemic regions marked by sustained human-to-human transmission often within sexual networks.^4^ Clade IIb emerged with sustained human transmission in Nigeria in 2014,^5,6^ driving the Nigerian epidemic that gave rise to the global 2022 multi-country outbreak.^7,8^ More recently, Clade Ib emerged in 2023, causing an outbreak in the eastern Democratic Republic of the Congo (DRC) that was strongly linked to dense sexual transmission networks.^9,10^ The viral lineages underlying these epidemics show strong APOBEC3-like mutational biases, a molecular signature characteristic of sustained human-to-human transmission.^5,6,9,11^

Sierra Leone confirmed its first mpox case in eight years on January 10, 2025, in a symptomatic adult male in the capital city, Freetown. A Public Health Emergency was declared shortly thereafter by the Ministry of Health.^12^ From January to July 2025, Sierra Leone rapidly became the epicentre of the continental mpox epidemic, accounting for nearly half of Africa’s confirmed cases during this period.^13^ By the 26th of September 2025, the country had recorded 5,328 confirmed cases and 56 mpox-associated deaths, with all 16 districts affected.^14^ Transmission was concentrated in the Western Area Urban and Western Area Rural districts, where Freetown’s dense and highly mobile population (∼1.4 million of the nation’s 8.6 million residents) facilitated inter-district spread that outpaced the deployment of interventions such as testing, case isolation, and contact tracing.

Preliminary genomic analysis indicate that the Sierra Leone outbreak dervises from Clade IIb/sh2017 lineage A.2.2 viruses circulating in Nigeria and neighboring West African countries.^15^ Epidemiological data show that the epidemic in Sierra Leone is predominantly affecting young adults aged 16-35.^12,16^ Unlike the early 2022 Clade IIb global wave and the 2017–2022 Nigerian epidemic, in which ∼63% of cases were male,^17,18^ the Sierra Leone outbreak exhibits a near-equal sex distribution, few pediatric cases, and a skew toward younger adult women and older adult men, patterns consistent with transmission within sexual networks.^12,16^ This demographic profile, together with the sharp increase in incidence observed around May 2025, more closely resembles the Clade Ib/sh2023 outbreaks reported in the DRC, Burundi, and Uganda.^10,19,20^ These similarities raise the possibility that contrasting demographic profiles across MPXV clades may reflect differences in transmission networks and contact dynamics rather than underlying viral genetic differences.^10^

The limited availability of full-length MPXV genomic data from Sierra Leone and neighbouring countries has left key questions about MPXV transmission dynamics in the region unresolved.^21^ In particular, it remains uncertain whether MPXV emerged locally in Sierra Leone and when and where this emergence occurred, as well as the mechanisms underlying its rapid nationwide spread. Here, we address these questions using comprehensive phylogenetic and phylogeographic analysis of new MPXV genomes sampled across Sierra Leone between January 10 and August 3, 2025. Understanding the spatiotemporal origins of the Sierra Leone epidemic lineage and its transmission dynamics is critical for guiding evidence-based response and strengthening preparedness for potential future outbreaks.

## Results

### Novel G.1 lineage drives the outbreak in Sierra Leone

To investigate the evolutionary relationship between Sierra Leone’s (SLE) epidemic lineage and global MPXV diversity, we generated 338 high-quality MPXV genomes from cases identified across the country between January 10 and August 3, 2025 (Figure 1A). The dataset was produced through collaboration among the Central Public Health Reference Laboratory (CPHRL, n=128 sequences), the Kenema Government Hospital (KGH) Viral Hemorrhagic Fever (VHF) Laboratory (n=105), and the Institut Pasteur de Dakar (IPD) mobile laboratory in Port-Loko (n=105). The dataset represents cases from 14 districts: Western Area Urban (n=129), Kenema (46), Port Loko (39), Bo (32), Bonthe (18), Bombali (16), Tonkolili (16), Kailahun (5), Western Area Rural (15), Kambia (4), Koinadugu (12), Falaba (3), Kono (1), and Pujehun (2) (Figure 1B). We also included two genomes from the Western Area Urban district sequenced by the Beijing Institute of Technology, bringing the total to 340 genomes.^15^ We analyzed only high-quality genomes (>60% coverage at 25x to 8100x read depth). This dataset represents 6.9% of all PCR-confirmed cases reported in Sierra Leone up to August 2025 (https://clt.npha.gov.sl/outbreak.aspx) (Figure 1A, B).

**Figure 1.**
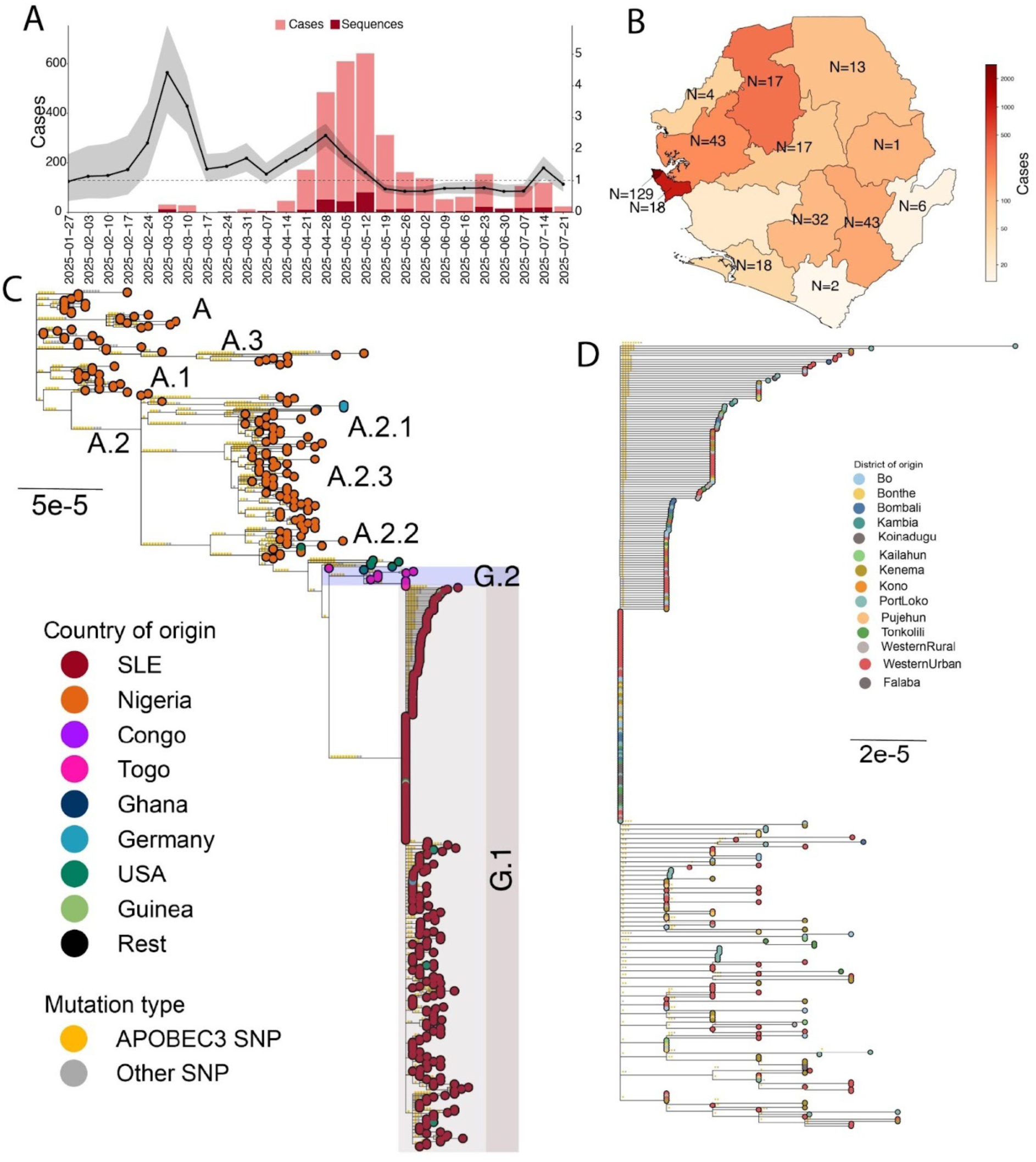
Phylogenetic origin and emergence of mpox lineage G.1 in Sierra Leone. A) Confirmed mpox incidence in Sierra Leone during 2025, showing the sequencing rate of this study over time (bars) and the time varying reproductive number (line, right y-axis, with95% confidence intervals). Annotations mark the start of vaccination campaigns in Western Area Urban (W.A.U.) and Western Area Rural (W.A.R.), as well as the launch of the national vaccination campaign. B) Number of confirmed mpox cases per district in Sierra Leone, with colour-coding by district as shown in the legend. The number of sequences generated per district in this study is annotated in the text. C) Clade IIb phylogeny with reconstructed single-nucleotide polymorphism (SNPs) mapped onto branches and colour-coded by country of sampling. We performed ancestral state reconstruction across our Clade IIb phylogeny to map SNPs to their corresponding branches. We annotated APOBEC3-characteristic substitutions (CT→TT or GA→AA), in the correct dimer context along branches and calculated their relative proportion across internal branches. APOBEC3 substitutions along the branches are shown in yellow, and all other substitutions in gray. Our new sequences are annotated in red and as G.1 in the text. The tree was rooted to the new zoonotic outgroup identified in (Parker et al. 2025). D) Focal G.1 lineage, with sequences colour-coded by sampling districts in Sierra Leone, as shown in the legend.

We reconstructed the Clade IIb phylogeny to determine the relationship between the SLE sequences and the Clade IIb/sh2017 lineage circulating in humans across West Africa. In our phylogeny, 339 of our 340 genomes clustered within the Clade IIb/sh2017 lineage, which emerged in humans in southern Nigeria in August 2014 and has since driven the ongoing human epidemic in West Africa.^5,6^ According to the nomenclature proposed by Happi *et al*., and Ruis *et al*.,^22,23^ Clade IIb/sh2017 is designated as Lineage A (or Clade IIb/sh2017/lineage A), with direct descendants designated as, e.g., A.1, and subsequent subdivisions as, e.g., A.1.1 - analogous to the Pango nomenclature used for SARS-CoV-2.^24^ Within Lineage A, our sequences form a well-supported monophyletic group (100% bootstrap support) descended from Lineage A.2.2 (Figure 1C). In accordance with the nomenclature, we designate the new SLE lineage as G.1, the alias of A.2.2.1, or Clade IIb/sh2017/lineage G.1 (Figure 1C).

Enrichment of APOBEC3 mutations in MPXV genomes serves as a molecular marker that distinguishes sustained human transmission from zoonotic spillover events.^6^ To confirm that the G.1 epidemic reflects sustained human-to-human transmission, we analysed our SLE genomes for APOBEC3-related mutational bias by reconstructing single-nucleotide polymorphisms (SNPs) across the phylogeny and quantifying the relative proportion of APOBEC3-like mutations. The G.1 lineage exhibits substantial APOBEC3-like mutational enrichment, with approximately 85% (90/106) of reconstructed SNPs consistent with APOBEC3 editing (Figure 1D). Together, these findings indicate that sustained human-to-human transmission of the newly identified Clade IIb/sh2025/lineage G.1 is driving the epidemic in Sierra Leone (Figure 1C-D).

We identified a single non–Clade IIb genome, a Clade IIa sequence sampled in mid-January 2025 in Western Area Urban. It clusters with two Clade IIa genomes from Guinea sampled in August and December 2024. Together, these three genomes form a lineage whose basal node corresponds to the 1965 Rotterdam zoo epizootic (KJ642614.1)^25^ and near-contemporary museum orangutan sequences,^26^ providing historical context for the Sierra Leone–Guinea cluster (Supplementary Figure 1). This clade lies downstream of the 1958 Copenhagen captive-monkey that marked the discovery of mpox,^27^ reinforcing that Clade IIa has long circulated in West Africa. Along the branch from the Rotterdam node toward Sierra Leone–Guinea, we observed 11 non-APOBEC3 substitutions and one APOBEC3-compatible change. The terminal end of the Sierra Leone genome carries 10 non-APOBEC3 and 1 APOBEC3-like substitution; whereas the Guinea genome from December 2024 have 5 non-APOBEC3 and 2 APOBEC3 changes; and the August 2024 Guinea genome shows just 2 non-APOBEC3 changes with no APOBEC3 events (Supplementary Figure 1). The relatively small number of APOBEC3 substitutions in these Sierra Leone—Guinea genomes, unlike the APOBEC-enriched Clade IIb sh/2017 G.1 lineage indicative of sustained human transmission, supports a zoonotic spillover with limited onward spread. However, we cannot yet determine whether the Sierra Leone case was imported from Guinea or represents an independent zoonotic event; additional sampling will be required to resolve this.

### G.1 descended from lineages circulating in West Africa

The geographic origin of the G.1 lineage remains unresolved. It is unclear whether it arose directly from endemic Clade IIb/sh2017 sublineages in Nigeria or was introduced into Sierra Leone via an intermediate source. In our phylogenetic analyses, G.1 is nested within Lineage A.2.2 diversity primarily sampled from the United States (U.S.; Figure 1C). G.1 is separated from its closest relative, a sequence from Togo, by 9 APOBEC-like mutations and two non-APOBEC3-like mutations along its stem branch. Previous studies have estimated that APOBEC3-like mutations accumulate at a rate of approximately 6 per year ^5,6^. The number of APOBEC3-like mutations along the G.1 stem branch suggests that it diverged from its ancestor, the A.2.2 sublineage, approximately eighteen months ago. This provides a lower bound on the timing of the lineage’s introduction into Sierra Leone.

The 11 A.2.2 genomes from the U.S. were sampled between June 2024 and June 2025 from Illinois (2), California, Massachusetts (2), Georgia, Pennsylvania, Tennessee, Minnesota, Michigan, and Virginia, with confirmed travel history to Nigeria for at least four of the sequences (Figure 1C). Based on the long internal branches separating the U.S. A.2.2 genomes, they most likely represent independent viral imports from the Nigerian A.2.2 lineage rather than an established lineage that has been cryptically diversifying in the U.S.. The closest publicly available Nigerian A.2.2 sequence to the A.2.2 lineage from which the USA and G.1 sequences descend is PP853012, sampled in Rivers State in September 2022. Taken together, these findings suggest that Lineage A.2.2 in the US and the G.1 lineage in Sierra Leone both descend from the Clade IIb/sh2017 lineage A.2.2, which originated in Nigeria and continues to seed localized epidemics outside the country. However, due to limited sampling in the region and the divergence time inferred from APOBEC3 mutations along G.1 stem branch, we cannot rule out the possibility of viral importation from an intermediate location.

Our phylogeny also indicates G.1 export from Sierra Leone (Figure 1C). This evidence includes four sequences sampled in the U.S. between March and June 2025, including three with confirmed travel histories to Sierra Leone. We also identified two sequences from Germany sampled from a single patient in March 2025, and two sequences from Guinea sampled in June 2025, with travel histories to Sierra Leone confirmed for all but the Guinea sequences. The detection of multiple viral export events indicates active local transmission in Sierra Leone at a prevalence sufficient to generate repeated international spread.

### G.1 emerged in late September 2024

The timescale inferred from accumulated APOBEC3-like mutations along the G.1 stem branch suggests that the lineage circulated undetected in Sierra Leone or in an unsampled external location for an extended period of time before detection. To estimate the timing of G.1’s emergence, we used Bayesian phylogenetic reconstructions in BEAST^28,29^ with a nested exponential model. Under this model, we apply an exponential growth model to the G.1 lineage while allowing the rest of the tree to evolve under an independent exponential growth model, capturing the distinct epidemiological dynamics observed in Sierra Leone.

We estimated that the time to the most recent common ancestor (tMRCA) of the G.1 lineage was 27 September 2024 [95% HPD: 12 August 2024 - 11 November 2024] (Figure 2A). This tMRCA represents the lower bound on when the sampled G.1 lineage became established in Sierra Leone. This suggests that the G.1 lineage may have circulated for approximately 3 months [95% HPD: 2 - 5 months] before its detection in early January 2025.

**Figure 2.**
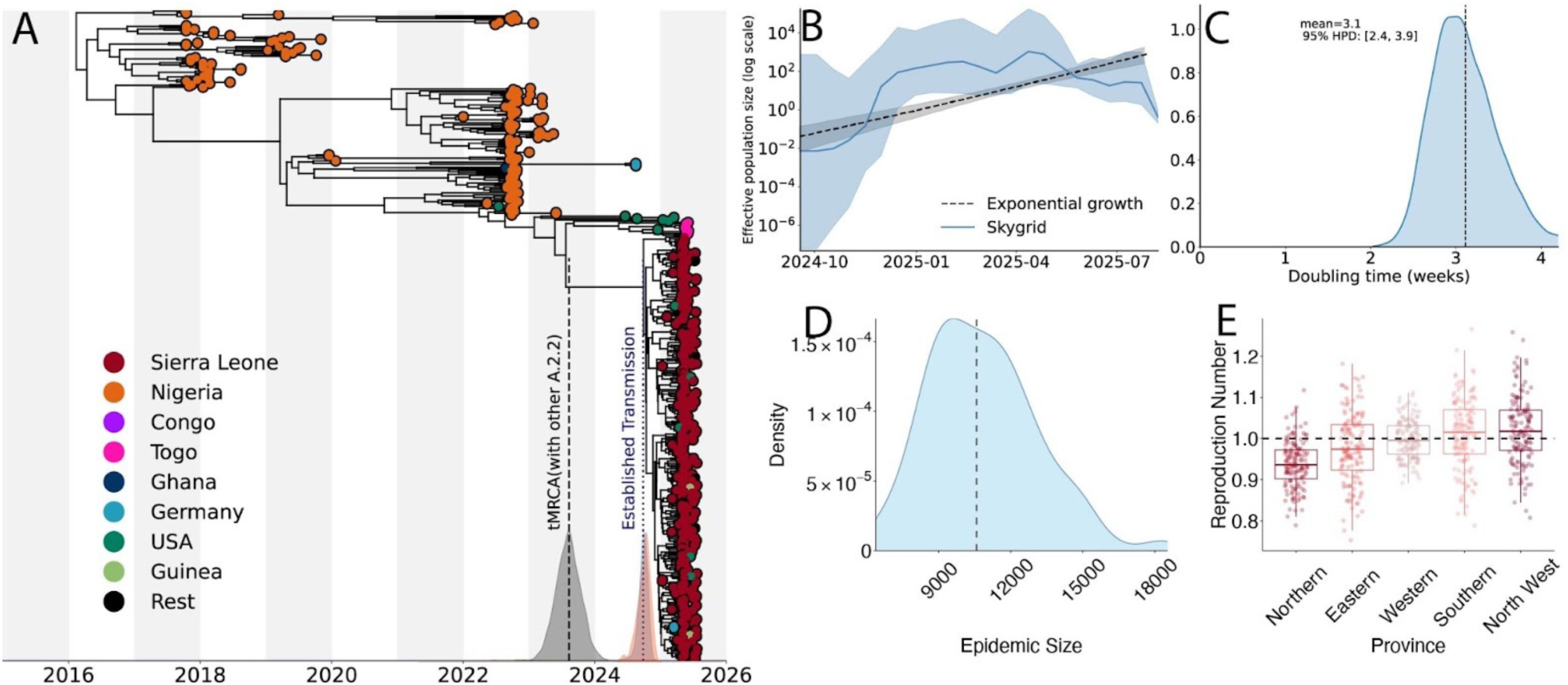
Timing, growth dynamics, and establishment of G.1 within Sierra Leone. A) Bayesian maximum clade credibility (MCC) tree of Clade IIb indicating when G.1 became established in Sierra Leone. Distributions on the x axis show 95% highest posterior density (HPD) intervals for three time estimates: 1) the tMRCA between G.1 and its closest A.2.2 lineage (gray; estimated by BEAST), representing the upper bound of the time of introduction into Sierra Leone 2) the tMRCA of the G.1 lineage (blue; BEAST), representing the lower bound on the time G.1 was established in Sierra Leone. 3) the epidemic start date inferred by JUNIPER (red) B). Effective population size through time for the Sierra Leone epidemic under a Skygrid (solid) and exponential (dashed) coalescent models. C) Posterior distribution of the estimated doubling time of the G.1 epidemic in Sierra Leone. D) Inferred epidemic size distribution from JUNIPER based on the estimated sampling proportion; the dashed line represents the mean. E) Boxplots of the reproduction number estimates per province; points denote the average reproduction number of all sequenced cases in each province across independent MCMC samples (see Methods); the dashed line represents unity.

In our exponential phylodynamic model, the inferred growth rate for the G.1 lineage corresponds to a doubling time of approximately 3 weeks [95% HPD: 2.4 – 3.9 weeks] (Figure 2B-C),consistent with the sharp increase in incidence observed between late April and May 2025.^16^ To confirm these observations, we estimated the time-varying reproductive number (Rt) from available case counts and obtained an Rt of 4.4 [95% CI: 3.1 - 5.9] in early March, consistent with both epidemiological data and the inferred doubling time (Figure 1A). ^30^ We also applied JUNIPER ^31^ to infer the transmission dynamics of the outbreak. JUNIPER estimated an outbreak start date of 11 September 2024 [95% HPD: 2 June 2025 - 11 November 2024], consistent with BEAST results, and an overall reproductive number of the outbreak, including the later decline, of 1.1 [95% HPD: 1.1 - 1.2]. In contrast, the estimated growth rate for the Nigerian epidemic, represented by the remainder of lineage A from which G.1 descended (Figure 2A), corresponded to a doubling time of approximately 2.6 years [95% HPD: 1.9 -3.4 years] (Supplementary figure 2). This is indicative of much slower growth in a more established epidemic. Our phylogenetic reconstruction under a non-parametric Skygrid coalescent model also supports a rapid and sustained increase in the effective population size of the G.1 lineage between April and July 2025 (Figure 2B). We additionally used the size of clusters of identical sequences to infer a similar reproductive number (1.2, 95% CI: 1.1 - 1.3) and an overdispersion parameter of 0.4 [95% CI: 0.2 - 1], consistent with previous reports of mpox transmission (Supplementary Figure 6).

Notably, in our JUNIPER analyses, we estimated that the sampling proportion (the fraction of cases sequenced) to be 3.3% [95% HPD: 2.3% - 4.8%]. This corresponds to an inferred total epidemic size of approximately 10,400 cases[95% HPD: 7,000 - 15,200], compared with 5,096 confirmed cases (Figure 2D). In addition, we estimated that the tMRCA between the G.1 lineage and the closest A.2.2.1 relative from Togo (i.e. the stem branch age) was August 9, 2023 [95% HPD: April 3, 2023, to December 19, 2023] (Figure 2A). This represents an upper bound on the time of the viral introduction into Sierra Leone. However, the rapid exponential growth observed and the absence of closely related A.2.2 samples from endemic Nigeria suggest that the introduction most likely occurred shortly before the tMRCA of G.1 in late September 2024.

### G.1 was established in the Western Area Urban district

The Western Area Urban and Rural districts are the epicentre of the outbreak, accounting for approximately 80% of the mpox cases in Sierra Leone. These districts include the capital, Freetown (∼1.4 million of the country’s 8.6 million residents; Figure 1B).^16^ However, it remains unclear whether the epidemic originated in this region or whether there was substantial under-ascertained transmission elsewhere during the early phase of the outbreak. To address these uncertainties, we performed discrete and continuous phylogeographic analyses at the district level to characterise the spatiotemporal spread of lineage G.1 within Sierra Leone.

Our discrete phylogeographic reconstructions indicate that lineage G.1 became firmly established in the Western Area Urban region, with strong posterior support (Posterior probability = 0.995; Figure 3A). This finding is consistent with the index case being reported from this district. However, the long stem branch of G.1 suggests an extended period of undetected circulation, potentially in an intermediate location, before detection in Sierra Leone. We therefore cannot confirm that Western Area Urban was the site of G.1’s initial emergence. Nevertheless, it is highly likely that G.1 became established there by late September 2024, with the district’s dense and highly mobile population driving onward spread across the country (Figure 3A).

**Figure 3:**
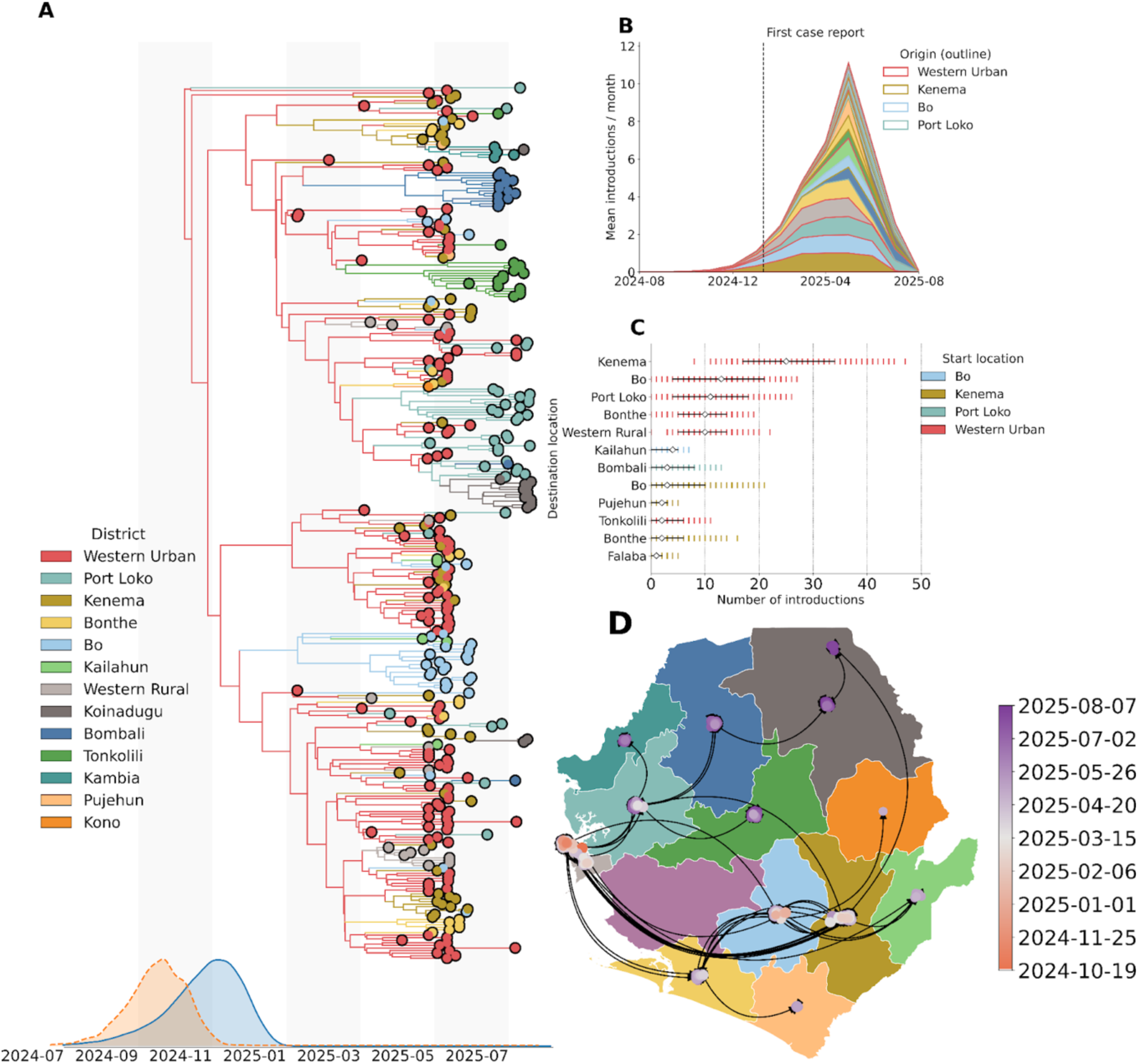
Spatiotemporal dynamics of mpox lineage G.1 in Sierra Leone. A) Discrete phylogeographic reconstruction showing the spatiotemporal spread of G.1 across Sierra Leone. Branches of the Maximum Clade Credibility tree (MCC) are coloured by source district, as indicated in the legend. The posterior distribution for the for the time to the most recent common ancestor (tMRCA) of the root of the tree (orange) and for the establishment of G.1 in Sierra Leone (blue) are shown along the x-axis. B) Distribution of the mean number of introductions per month by district. Each bar represents the end location of an introduction is coloured by districts as in panel A the origin district of each introduction is outlined according to the legend in panel B. C) Total number of introductions between districts, shown as coloured bars representing the original district (x-axis) and destination district (y-axis), with 95% highest posterior density (HPD) intervals indicated in black D) Continuous phylogeography of G.1 spatiotemporal spread across Sierra Leone, with timing of viral dissemination indicated by the colour gradient as in the legend. The inferred movement of the epidemic proceeds in an anticlockwise direction. The boundary data for the map is from GADM (http://www.gadm.org).

To investigate this pattern of within-country spatiotemporal spread while accounting for uncertainty in phylogeographic reconstruction due to sparse sampling, we incorporated a Markov jump counting approach to estimate the timing and origin of geographic transitions across the full posterior distribution at both regional and district levels. The analysis confirmed that the Western Area Urban was the primary source of interdistrict viral dissemination throughout the outbreak, with an estimated 71 introductions [95% HPD: 56 - 88] originating from the district (Figure 3B). Normalised by its 2021 population (609,174), this corresponds to ∼117 exports per million residents (HPD 92–144 per million), far exceeding any other district.

Most early viral exports originated in Western Area Urban region and spread into Kenema, followed by Bo, Port Loko, Western Area Rural, and Bonthe (Figure 3B, C). Although Western Area Urban remained the principal source throughout the epidemic, Kenema became an important secondary hub later in the outbreak, accounting for an estimated 10 exports [95% HPD: 3 to 21], equivalent to ∼13 per million (HPD 4–27 per million; population 772,472). with exports predominantly disseminating to Bo, Pujehun, Bonthe, and Falaba (Figure 3C). The remaining districts contributed only marginally to viral dissemination. Continuous phylogeography analysis was consistent with these discrete analyses, indicating that G.1 was first established in the Western Area Urban region before spreading early into Western Area Rural, Kenema and Bonthe (Figure 3D).

In our dataset, Port-Loko was disproportionately well sequenced relative to other regions, including the Western Area Urban, with 21% of confirmed cases sequenced compared to 5% in the Western Area Urban. To account for this heterogeneity in sampling across districts, we repeated the discrete phylogeographic analysis at the regional level. The regional reconstructions were consistent with the district-level results, again indicating that G.1 most likely emerged or was first established in the Western region (posterior probability=0.99) (Supplementary Figure 3).

Our regional analyses also supported the patterns of within-country spatiotemporal spread observed in the district-level analyses. We found that the Western region was the principal source of inter-region viral introductions, with an estimated 52 introductions [95% HPD: 39 to 67], equivalent to ∼41 introductions per million residents (95% HPD, 31–53 per million). The earliest introductions originated in the Western region and disseminated to the Eastern, Southern, and Northwest regions (Supplementary Figure 4-5). As the epidemic progressed, the introduction profile shifted, with subsequent spread into other regions increasingly driven by viral exports from the Western, Eastern, and Southern regions (Supplementary Figure 5). The Northern regions contributed only marginally to inter-district spread, with transmission there primarily seeded by introductions from the Western region (Supplementary Figure 5).

### Persistence in the Western Area Urban drove the outbreak

As shown above, repeated introductions from a limited number of districts drove the spread of lineage G.1 across Sierra Leone. However, the extent to which locally persistent transmission chains sustained ongoing transmission within districts remained unclear. To distentangle the relative contributions of local viral persistence versus new introductions, we quantified the duration of within-district viral persistence for each reconstructed transmission chain (Figure 3A, 4A).

We found evidence of persistent within-district circulation in the Western Area Urban district from the time of emergence onwards (Figure 4A). A large transmission chain originating in this district persisted continuously throughout the sampling period (Figure 4A). We also observed an early introduction from the Western Area Urban into Port Loko in November 2024, with the resulting transmission chain sustaining local circulation for the duration of the epidemic, detectable despite sparse sampling (Figure 4A). This pattern is consistent with the index case’s recent travel history to Lungi, located in the Port-Loko district and home to the country’s international airport ^15,32^. By the time the first cases were detected in early January 2025, active transmission chains had already been established in four districts: Western Area Urban, Port-Loko, Bo, and Kenema (Figure 4A). Several districts, including Bo, Bombali, and Kenema, harboured transmission chains seeded from Western Area Urban in early 2025 that persisted locally for months (Figure 4A, B).

**Figure 4.**
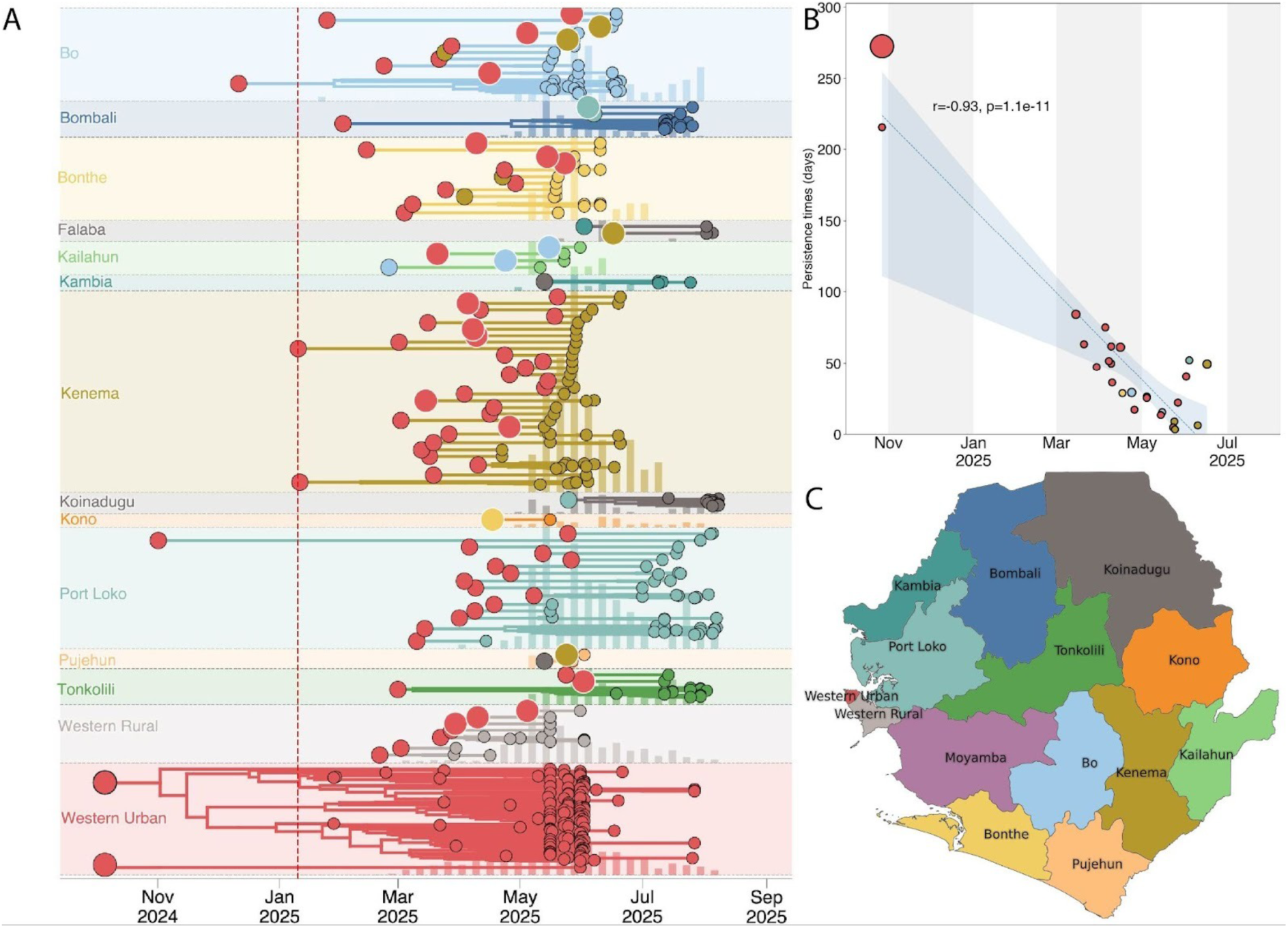
Persistence and local transmission dynamics of G.1 across Sierra Leone. A.) Duration and timing of district-level transmission chains inferred from the phylogenetic reconstruction. Each coloured band represents a distinct transmission chain grouped by district (background colour as in panel C). The circle at the origin of each chain denotes the district of origin, with size proportional to the number of descendant tips in that clade. Grey vertical ticks behind each band show weekly reported case counts for that district, rescaled to band height for visual context. The red dashed line marks a date of the first reported case in Sierra Leone (10 January 2025). B) Relationship between clade persistence (days) and clade origin time, with oints coloured by district as in panel A. The fitted line shows the least-squares regression with 95% confidence intervals. A strong negative correlation was observed (Pearson r = −0.93, p = 1.1×10⁻¹¹), indicating shorter transmission chain persistence later in the outbreak. C) Map of Sierra Leone district locations, coloured consistently with panels A-B. The boundary data for the map is from GADM (http://www.gadm.org).

From March 2025 onward, the number of transmission chains co-circulating within and across districts increased sharply, consistent with the rise in incidence observed between late April and May 2025 (Figure 4A, 1A). These transmission chains were predominantly seeded from the Western Area Urban district, with repeated introductions establishing multiple co-circulating chains with comparatively shorter periods of persistence in districts such as Kenema, Western Area Rural, and Bonthe (Figure 4A). Although our sampling period limits inference beyond August 2025, the data suggest that sustained viral persistence, driven by undetected or mild infections, maintained transmission in the Western Area Urban district, while local epidemics in other districts were primarily fueled by repeated introductions and transient transmission chains from this dominant source.

## Discussion

Mpox remains a significant and evolving public health threat, with both Clade I and Clade II now demonstrating sustained human transmission and driving large-scale outbreaks across Africa and beyond.^5–10^ Since its emergence in 2014, Clade IIb has persisted in human populations, fueling both the global multi-country B.1 outbreak in 2022, and now the G.1 outbreak in Sierra Leone.^5,6,8,33^ This shift toward sustained human-to-human transmission, observed independently for Clade Ia, Clade Ib, and Clade IIb, appears driven primarily by epidemiological factors, such as transmission in dense sexual networks, though a contribution from viral fitness cannot be ruled out. This study and others found no evidence of APOBEC3-mediated evolution that would result in increased transmissibility, supporting the view that recent expansions have been driven primarily by human behavioral and network factors..^5,6,10,33^

Our phylogeographic analyses indicate that G.1 was most likely established in the Western Area Urban district in Sierra Leone, which served as the primary and persistent source for viral dissemination across Sierra Leone. Although early cases were concentrated within sexual transmission networks, subsequent spread became more generalised, maintained through both household and community contact transmission alongside sexual networks, a pattern paralleling recentClade Ib in DRC, Burundi, and Uganda.^12,16^ Taken together, these findings suggest that the rapid expansion of G.1 from April to May 2025, was driven primarily by population dynamics and network connectivity, even as the potential influence of viral adaptation remains uncertain. This underscores that effective epidemic control remains achievable through targeted interventions that disrupt key transmission networks.

Our findings also underscore the urgent need to strengthen early warning surveillance systems and expand access to diagnostics for outbreak control across Sierra Leone. We estimate that lineage G.1 cryptically circulated for approximately three months before detection, spreading to at least three districts beyond Western Area Urban district during this period. The true epidemic size was estimated to be 10,400 cases (with 5,096 laboratory-confirmed cases), indicating that surveillance systems did, however, detect approximately 49% of total cases across the outbreak. Strengthening national surveillance capacity, particularly for early detection will require proactive, community-based case finding, decentralised diagnostics with stable supply chains,; and real-time genomic sequencing.

At the same time, strengthening frontline capacity is essential to translate surveillance data into effective outbreak control. Training of healthcare workers and community health volunteers should emphasise recognition of mpox’s early and distinguishing signs, and its differentiation from varicella where both infections are endemic.^34–36^ It should reinforce adherence to standardised case definitions, safe specimen collection, and rigorous infection-prevention and control (IPC) measures including isolation, hand hygiene, personal protective equipment (PPE) use, and safe caregiving. In addition, risk-reduction counselling and practical guidance are needed to help communities limit spread in households, schools, markets, and healthcare facilities. These investments will improve detection and case management not only for mpox but also for other epidemic-prone diseases^32^.

Mpox incidence in Sierra Leone peaked during weeks 18–19 (late April to mid-May), then began a steady decline as coordinated response measures took effect. By 17 August 2025, 147,779 individuals had been vaccinated, 51% of which were male and 83% aged 20–49 years. Vaccination coverage included health-care workers (23%), identified contacts (43%), high-risk groups (12%), and others (22%). Despite this modest national vaccination coverage of ∼1.96%, the timing of case decline aligns closely with key public health intervention strategies, including vaccination campaigns in the Western Area Urban and Rural districts in week 13, the nationwide vaccination campaign in week 18, immediate vaccination of high-risk contacts in week 20, ring vaccination of contacts in week 23, and the Enhanced Integrated mpox Response in week 26. Together, these actions underscore the importance of layered, timely vaccination strategies, supported by robust surveillance, in controlling transmission. Ensuring equitable and rapid vaccine access remains essential not only to reduce morbidity and mortality within Sierra Leone, but also to prevent regional spread of the outbreak.

This study has several limitations, primarily related to sampling coverage and spatial heterogeneity. Although our dataset includes ∼6.9% of confirmed cases, per-district sampling varied widely across districts; from fewer than 5% of cases in Western Area Urban (131 of 2,867) and Western Area Rural (15 of 1,020) to more than 30% in Kenema, Bo, and Bonthe.. Such uneven representation may bias phylogeographic inferences, overestimating persistence or exports from better-sampled districts while underestimating others. To mitigate this, we performed regional-level analyses that produced consistent patterns, though some residual bias likely remains. Our time-varying reproductive number estimates are further constrained by under-ascertainment, as suggested by the JUNIPER-estimated epidemic size of ∼10,400 cases compared with the 5,096 confirmed by qPCR. Nevertheless, key phylogeographic inferences, Western Area Urban as the primary source of spread with early seeding of Kenema, Bo, Port Loko, Western Area Rural, and Bonthe, are well supported by independent epidemiological patterns in the case data (largest absolute burden in Western districts, subsequent growth in Kenema, Bo, and Port Loko). Finally, the lack of detailed clinical and behavioural metadata prevents direct linkage of genomic patterns to disease severity, viral load, or transmission context.

This study highlights both the power of sustained investment in detection and sequencing. The rapid identification of the G.1 lineage and its spread across Sierra Leone demonstrates how local capacity, when paired with timely genomic data, can transform outbreak response from reactive to informed. Yet it also underscores how fragile these gains remain: without equitable access to diagnostics, vaccines, and therapeutics, mpox and other zoonotic pathogens will continue to emerge and sustain transmission across Africa. If not contained, these pathogens could escalate into global health threats. To prevent this, we must move beyond crisis-driven responses that depend on external aid toward long-term preparedness and resilience, grounded in global solidarity, sustained investment including domestic funding, and strengthened local health systems.^37^ Expanding real-time sequencing, representative surveillance, integrated clinical–genomic data systems, and research capacity will be critical to ensuring that early detection translates into effective control, not only for mpox but also for future emerging pathogens.

## Methods

### Ethics statement

This work was conducted as part of the public health response to the mpox outbreak under the mandate of the National Public Health Agency (NPHA), Ministry of Health, Sierra Leone. All activities were performed in accordance with national outbreak investigation guidelines. As this investigation formed part of an emergency public health response, it was exempt from institutional ethics review and the requirement for individual informed consent under national public health regulations. All data were anonymized before analysis to protect patient confidentiality.

### Sample collection and molecular testing

All samples were collected between 10th January and 3rd August 2025 from outbreak response surveillance. The dataset was produced through collaborative efforts between the Central Public Health Reference Laboratory (CPHRL), Kenema Government Hospital (KGH) Viral Hemorrhagic Fever (VHF) Laboratory, and the Institut Pasteur de Dakar (IPD) mobile laboratory in Port-Loko. Samples were processed for Deoxyribonucleic Acid (DNA) extraction at CPHRL and KGH VHF lab, using the RADI FAST DNA Extraction Kit (KH Medical, Seoul, South Korea), following the manufacturer’s standardized protocols. The extracted DNA was then tested for the presence of Monkeypox virus (MPXV) using the RADI FAST mpox Detection Kit via real-time quantitative PCR (qPCR), following the manufacturer’s instructions. At the IPD/Port-Loko Lab, DNA extraction was performed using the QIAamp DNA mini kit (50), following the manufacturer’s instructions ( Qiagen, Berlin, Germany), and qPCR was done using the Lyophilized 1-step RT-PCR Polymerase Mix kit with the Lightmix modularDx Monkeypox virus assay, according to the manufacturer’s instructions (Roche, Tibmol Biol, Berlin, Germany). Every experiment included appropriate internal and external quality controls.

### Next-generation sequencing

We prioritized mpox-positive samples with a cycle threshold (Ct) value ≤ 30 for sequencing. Libraries were prepared using the Illumina RNA Prep with Enrichment (L) Kit and enriched with the Viral Surveillance Panel (VSP) 2.0 targeting epidemic-prone pathogens, including MPXV. Following capture with streptavidin-coated magnetic beads, libraries were amplified, purified, quantified, and quality-checked using Qubit (Thermo Fisher Scientific) and Agilent TapeStation (Agilent Technologies, Santa Clara, CA, USA). Samples from CPHRL were sequenced on the Illumina MiniSeq with 2 × 150 bp paired-end reads. Sequences from KGH were generated using the Illumina VSP panel, followed by Illumina sequencing on Illumina Miseq. Sequences at the IPD mobile lab were generated using an amplicon-based method with specific mpox primer pools, followed by Oxford nanopore MK1B or Illumina iSeq100 sequencing ^38^.

### Genome assembly

The CPHRL sequences were generated with an in-house reference-based assembly pipeline. Briefly, we mapped reads against a Clade IIb reference genome (NC_063383, an early Clade IIb/sh2017 genome from Nigeria) with bwa-mem ^39^ and called consensus using samtools ^40^ and iVar ^41^. Sequences from KGH were assembled using the Broad Institute viral-ngs assemble_ref_based pipeline implemented in Terra (refs) using a Clade IIb reference genome (NC_063383). Sequences from IPD were assembled from Oxford nanopore data, using an in-house reference-based assembly pipeline with a Clade IIb reference genome (NC_063383) and a minimum read depth of 100. For illumina data, the trimmed BAM files were mapped to a Clade IIb reference genome (NC_063383) using BWA-MEM (version 0.7.17-r1188)^39^. The generated BAM mapping files were sorted using SAMtools (version 1.6) ^40^ and then used as input to iVar (version 1.3.1) ^41^ with a minimum read depth of 10 and depth threshold of 0.5, to generate consensus sequences. The mean coverage across all sequences ranged from 25x to 8100x, with 324 of the 338 genomes exceeding 70% completeness.

In total, we generated 338 high-quality sequences. The total sequences produced were: Central Public Health Reference Laboratory (CPHRL, n=128 sequences), Kenema Government Hospital (KGH) Viral Hemorrhagic Fever (VHF) Laboratory ( n=105), and the Institut Pasteur de Dakar (IPD) mobile laboratory in Port-Loko (n=105). Our dataset included sequences sampled between 10 January and 3 August 2025 from 14 districts: Western Area Urban (n=129), Kenema (46), Port Loko (39), Bo (32), Bonthe (18), Bombali (16), Tonkolili (16), Kailahun (5), Western Area Rural (15), Kambia (4), Koinadugu (12), Falaba (3), Kono (1), Pujehun (2).

### Genomic dataset curation

We combined our 338 genomes with all high-quality (genome coverage >70%) publicly available Clade IIb MPXV genomes from Pathoplexus.^42^ Only one representative of the multi-country outbreak lineage B.1 was included, as it was not the primary focus of our study. We also included the closest zoonotic outgroup to Clade IIb (PP852949.1) sampled in 2022 from Nigeria, which was used to root the tree.^5^ In total, the dataset comprises 530 sequences.

### Epidemiological data

We obtained the time series of confirmed mpox cases from the National Public Health Agency. We used EpiFilter/Smooth to infer a time-varying Rt.^30^

### Phylogenetic analysis

We aligned our dataset to the Clade IIb reference genome (NC_063383) using the squirrel package developed by O’Toole et al., ^6,43^. The alignment was trimmed, and the 3′ terminal repeat regions, as well as repetitive regions, regions with low complexity, clustered mutations or mutations near gaps or ambiguities, were masked using the package’s quality control mode.

The full MPXV phylogeny was reconstructed with IQ-TREE v2.0 implemented within the squirrel package.^44^ We reconstructed a separate Clade IIb phylogeny under the default parameters in Squirrel, and rooted it to the zoonotic outgroup PP852949 from Nigeria 2022 in Nigeria ^5^, which was subsequently pruned. All zero-length branches were collapsed, and an ancestral state reconstruction was performed across the Clade IIb phylogeny using IQ-TREE2.^44^. Lineages were assigned from the nomenclature established by ^22,23^ using the Nextclade tool.^45^

### Bayesian phylodynamics

To estimate the timing of G.1’s emergence, we adopted the partitioned model developed by O’Toole et al.^6^ to model APOBEC3-mediated evolution implemented in the BEAST X v10.5.0 software package^28,29^ with the BEAGLE high-performance computing library.^46^ We used a nested exponential coalescent model to adequately model G.1’s distinct epidemiological dynamics. We applied an exponential growth model to the tree from the most recent common ancestor (MRCA) of the G.1 lineage onwards. The rest of the tree, representing the remainder of Lineage A i.e. the source population, was modelled under an independent exponential growth model. This allows us to estimate distinct growth rates and associated doubling times for the epidemic in Sierra Leone and the wider West Africa (predominantly Nigeria) respectively. We ran two independent chains of 100 million states to ensure convergence, discarding the initial 10% of each chain as burn-in. The chains were then combined using LogCombiner. For all subsequent analyses, we assessed convergence using Tracer, and constructed a maximum clade credibility (MCC) tree in TreeAnnotator 1.10 ^47^.

#### Phylogeographic analysis

To investigate the spatial spread of the G.1 lineage within Sierra Leone, we reconstructed the timing and pattern of geographic transitions between Sierra Leone’s districts and at the regional level under an asymmetric discrete trait analysis.^48^ We applied Bayesian Stochastic Search Variable Selection (BSSVS) to identify statistically supported migration routes. We used a Skygrid coalescent model with 34 change points distributed over seven months to capture the doubling time of two weeks interval, excluding Lineage A sequences from other countries.^49^ We combined two independent MCMC chains of 100 million states sampled every 10000 states, discarding the initial 10% of trees as burn-in. We confirmed that all effective sample size (ESS) values were above 200.

We employed a Markov jump counting procedure across the full posterior distribution to further investigate the timing and origin of geographic transitions while accounting for uncertainty in our phylogeographic reconstructions. We used the TreeMarkovJumpHistoryAnalyzer implemented in BEAST X^28,29^ to extract all Markov jumps from our posterior tree distributions.^50^ We accounted for the uneven distribution of sequences across Sierra Leone’s districts by performing all phylogeographic analyses on an aggregated, regional level.

We also performed a continuous phylogeographic analysis to quantify the dispersal of the G.1 lineage across regions and districts in Sierra Leone. We assigned each sequence the latitude and longitude of the centroid of its sampling district. We used the Skygrid coalescent model described above, with a Cauchy distribution to model the among-branch heterogeneity in dispersal velocity.^51^ We ran two independent MCMC chains of 50 million states, sampling every 2000 states. We combined the chains after discarding 10% of the states as burn-in.

### Transmission reconstruction

We used the R package JUNIPER ^31^ to conduct outbreak reconstruction from all Clade IIb G.1 samples based on pathogen genomes. JUNIPER infers transmission networks using both sequenced and unsequenced cases. We masked our Clade IIb alignment only to include the APOBEC3 target site partition. Specifically, as APOBEC3 proteins mutate TC dimers to TT dimers and GA dimers to AA dimers, we retained only pairs of positions on the aligned genomes that exhibited TC or TT in all sequences, or GA or AA in all sequences. We removed the conserved nucleotide in each dimer across all sequences, to retain only the second nucleotide of the TC/TT dimers and retained only the first nucleotide of the GA/AA dimers. A substitution rate of 6.9e-7 was calculated by assuming 6 APOBEC3-like mutations per site per year across the partitioned genome length of 23,602.^5,6^ JUNIPER was run for a total of 30,000 iterations (across two 15,000 iteration runs) with a fixed mutation rate, a generation interval of 11.4 days, and a sojourn interval of 15 days. After completion of the runs, 20% burn-in was removed, and the resulting chains were downsampled to an effective sample size of 69 and 73, respectively, to ensure minimal autocorrelation between samples ^52^ and combined for a total ESS of 142.

The estimated outbreak size and estimated outbreak start date were directly inferred from the results. To calculate the per-individual reproductive number, we computed the expected number of total transmissions under the stochastic-epidemic model. The probability P(X = *k*) that an individual infected at time *t* transmits *k* total people, given that *d* offspring are explicitly represented in the transmission network inferred by JUNIPER, is given by

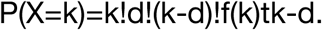

where ω^t^ is the probability that a case infected at time *t* has no sequenced descendants up through the time of last sample collection, and *f*(*k*) is the probability of one person transmitting to *k* other people. The equation is adapted from TransPhylo ^53^. The expected number of transmissions for said individual (implicitly conditional on them infecting at least d individuals) is then given by

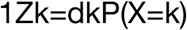

where

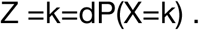

Reproductive numbers within any metadata category were computed by averaging the expected number of transmissions for each observed individual in said category.

## Data availability

All sequences are available on Pathoplexus (https://doi.org/10.62599/PP_SS_232.1). All other data are available at GitHub (https://github.com/Ifeanyi-omah/Mpox_project_sierraLeone)

## Code availability

All code to run the analyses is available at GitHub (https://github.com/Ifeanyi-omah/Sierra_Leone_Mpox_project)

## Funding Statement

Africa CDC, through the Africa PGI supported the establishment of in-country sequencing capacity at CPHRL through the provision of sequencing equipment, training, testing and sequencing reagents and the World Health Organization (WHO) provided also critical support through the provision of sample storage equipment, diagnostic and sequencing reagent supplies, complemented by essential training at the National level. This work is also made possible by support from Flu Lab and a cohort of generous donors through TED’s Audacious Project, including the ELMA Foundation, MacKenzie Scott, the Skoll Foundation, and Open Philanthropy. This work of the Institut Pasteur de Dakar (IPD) team in terms of sample testing, sequencing, capacity building, data analysis and dashboarding, and coordination, was funded by West African Health Organization (WAHO/OOAS), in support of the Sierra Leone national mpox response efforts. I.F.O. is supported by the Wellcome Trust Hosts, Pathogens & Global Health programme (Wellcome Trust, grant 218471/Z/19/Z) in partnership with Tackling Infectious Disease to Benefit Africa. A.R. acknowledges the support of the Wellcome Trust through the ARTIC Network (awards 206298/Z/17/Z & 313694/Z/24/Z).

## Acknowledgements

The Institut Pasteur de Dakar acknowledges Mamadou Cissé data scientist, and Mame Diarra lab technician, deployed with the IPD team in Sierra Leone, and Ndongo Dia, Oumar Faye, Cheikh Loucoubar, Diom Yero Sow and Ousmane Faye for logistics and coordination support from IPD. The IPD would also like to thank Landry Boussiengui and Fatou Diène Thiaw for generating the first sequences from Sierra Leone at IPD.

## Supplementary figures

**Supplementary Figure 1:**
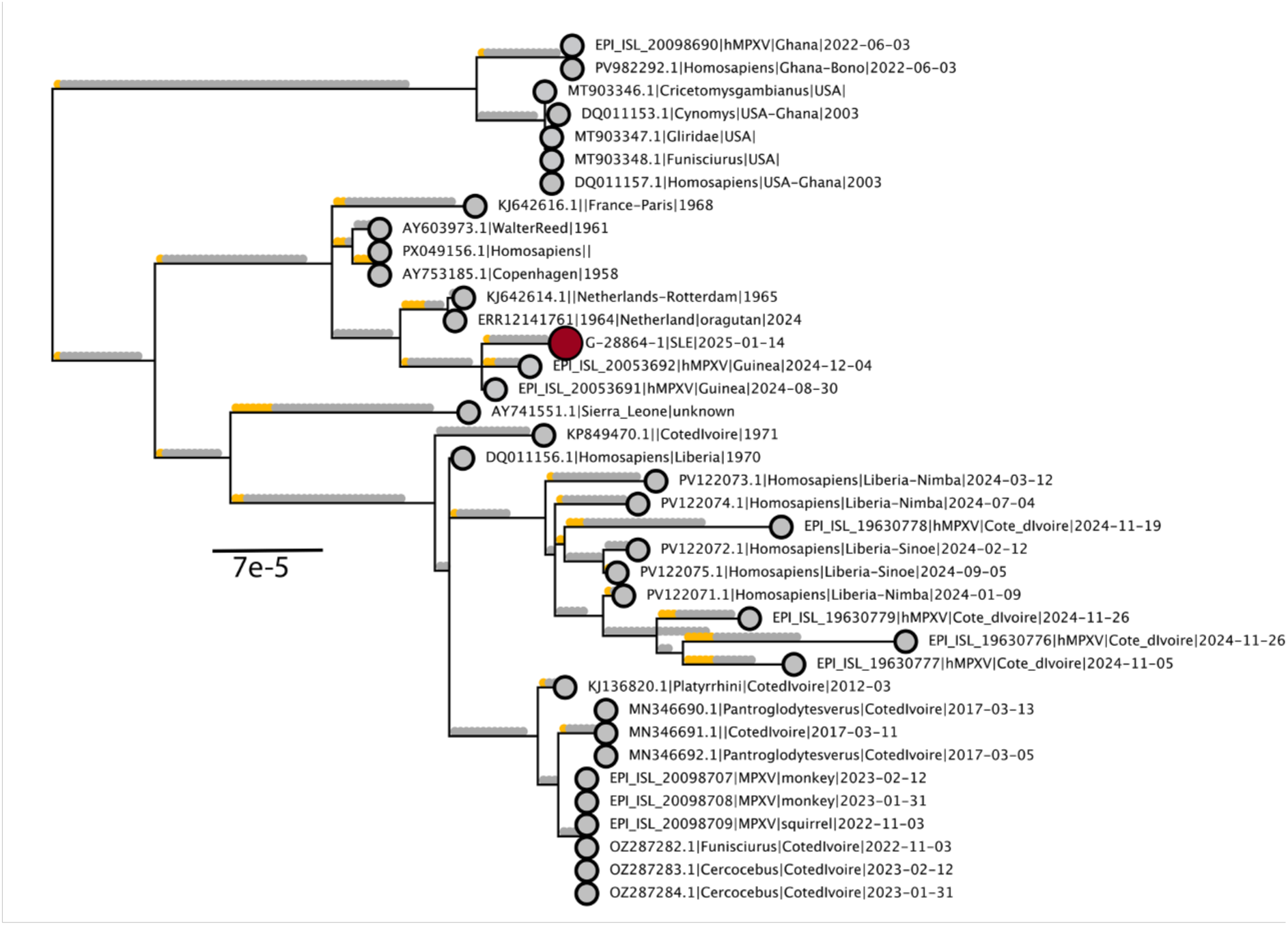
Clade IIa phylogeny with reconstructed SNPs mapped onto branches. We performed ancestral state reconstruction across our Clade IIa phylogeny to map SNPs to their relevant branches. We annotated APOBEC3 characteristic substitutions, i.e. CT or GA in the correct dimer context along branches and calculated their relative proportion across internal branches. APOBEC3 substitutions along the branches are annotated in yellow with the remainder in gray. Our new sequence is annotated in red as an enlarged tip. The tree was rooted to the new Clade IIb zoonotic outgroup identified in ^5^.

**Supplementary Figure 2:**
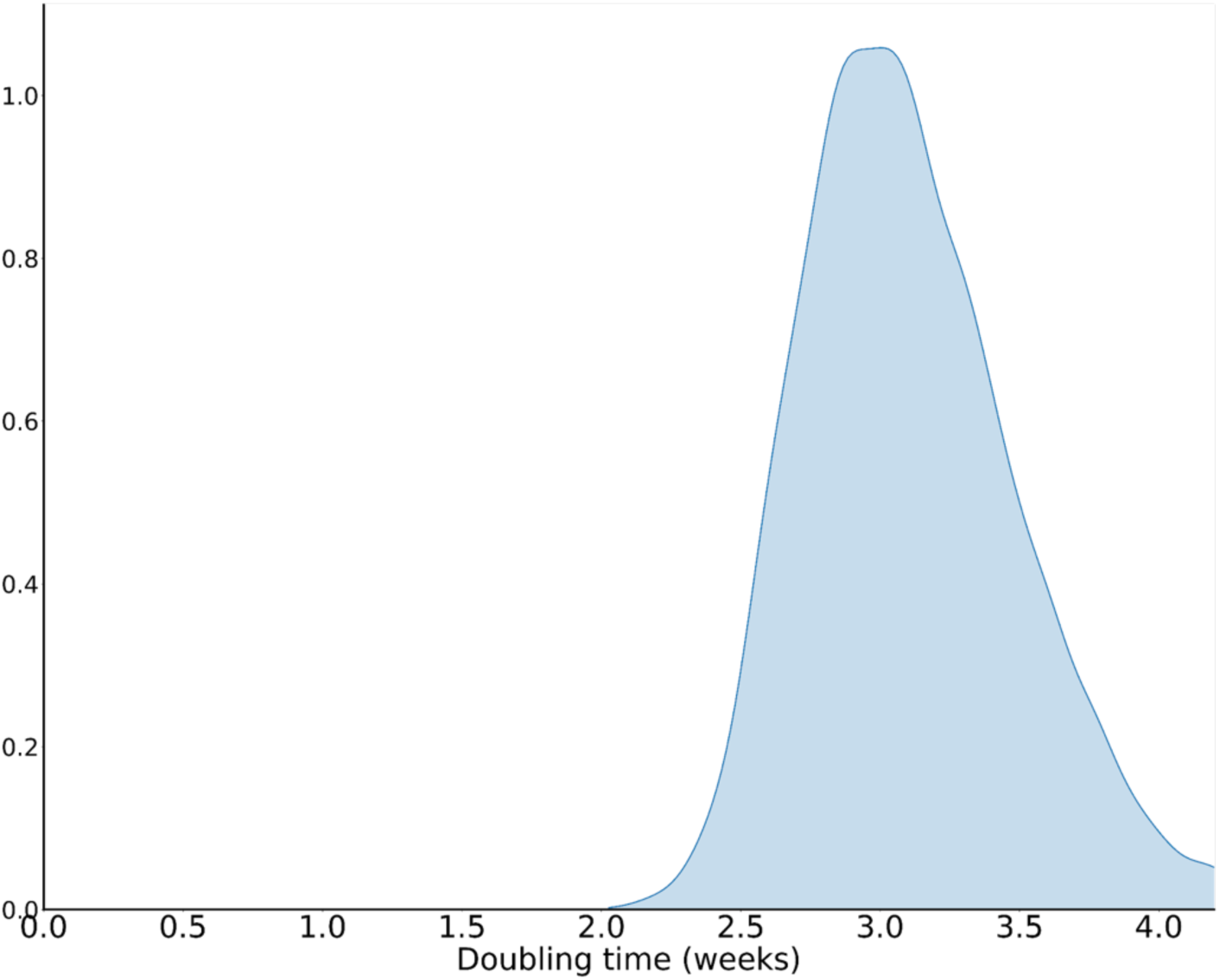
The distribution of the doubling time for the Nigerian epidemic from 2017 to 2022.

**Supplementary Figure 3:**
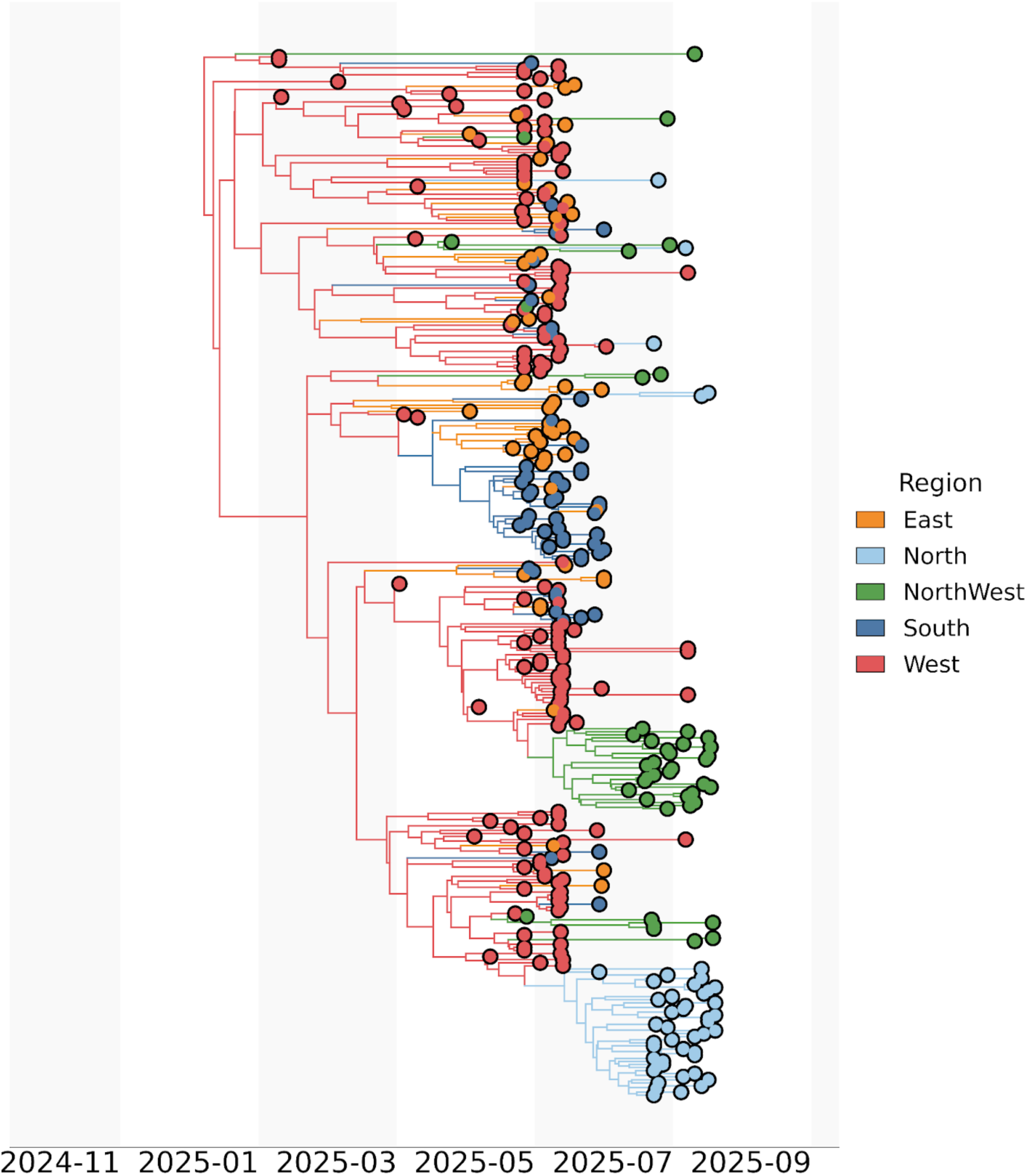
Phylogeographic analyses of the G.1 lineage in Sierra Leone on a regional level. The branches of the Maximum Clade Credibility tree (MCC) are coloured by region, as per legend.

**Supplementary Figure 4:**
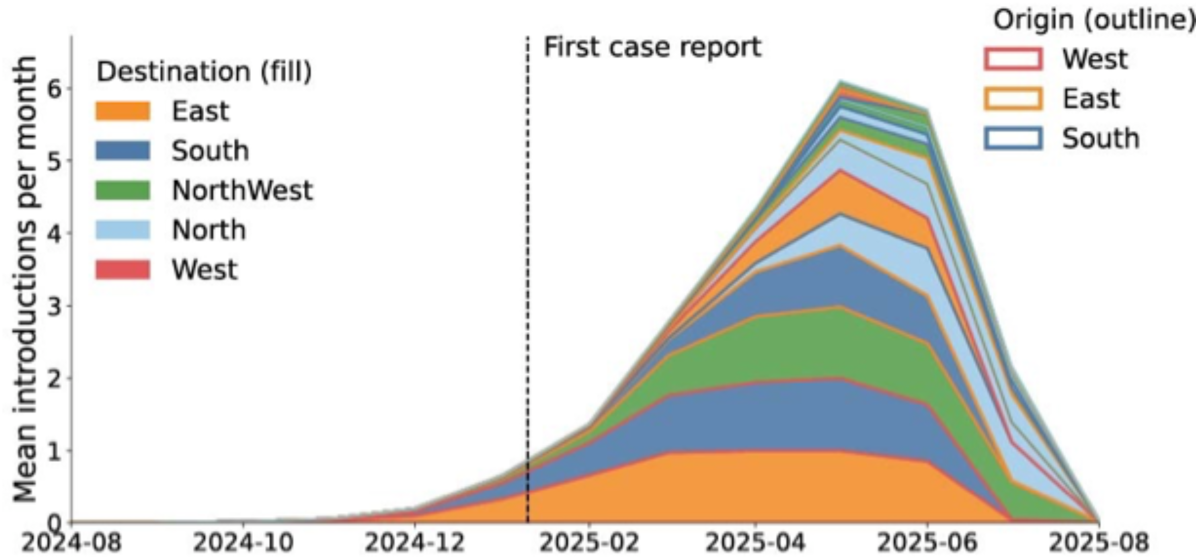
The distribution of the number of introductions across time by region. Data are summarised across the posterior of 10,000 trees. Distribution represents the 95% highest posterior density. The end location state is coloured by region, as per legend. The start location is highlighted by an outline of the area.

**Supplementary figure 5:**
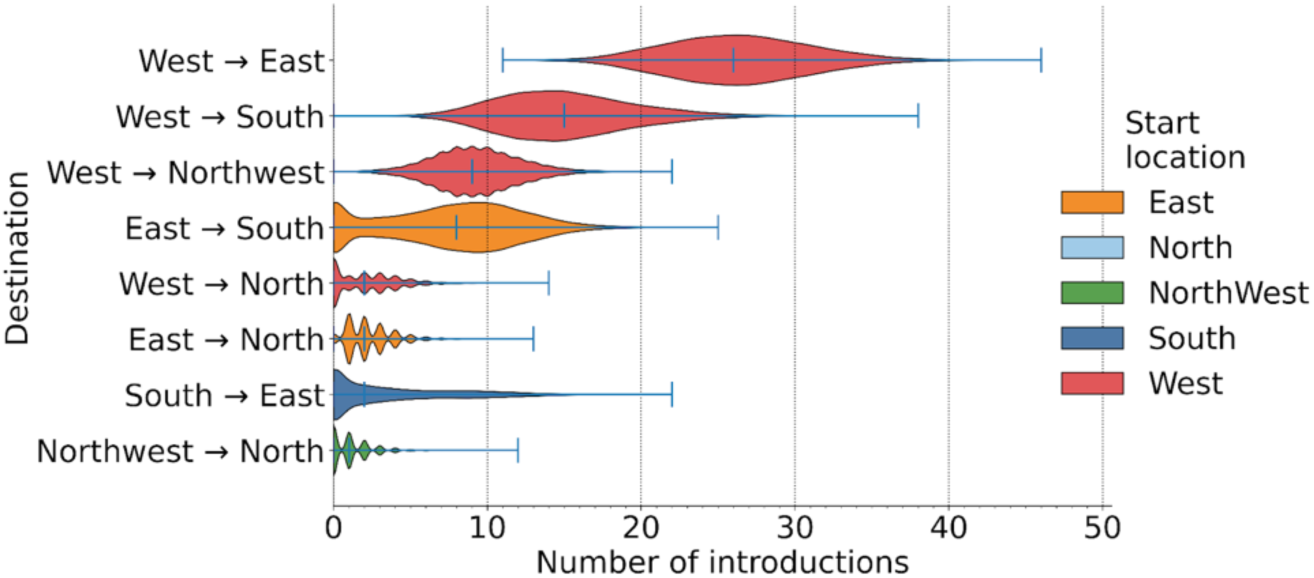
Total number of introductions by region from each start location. Data are summarised across the posterior, annotated as per legend in colour, to each end location on the y-axis.

**Supplementary figure 6.**
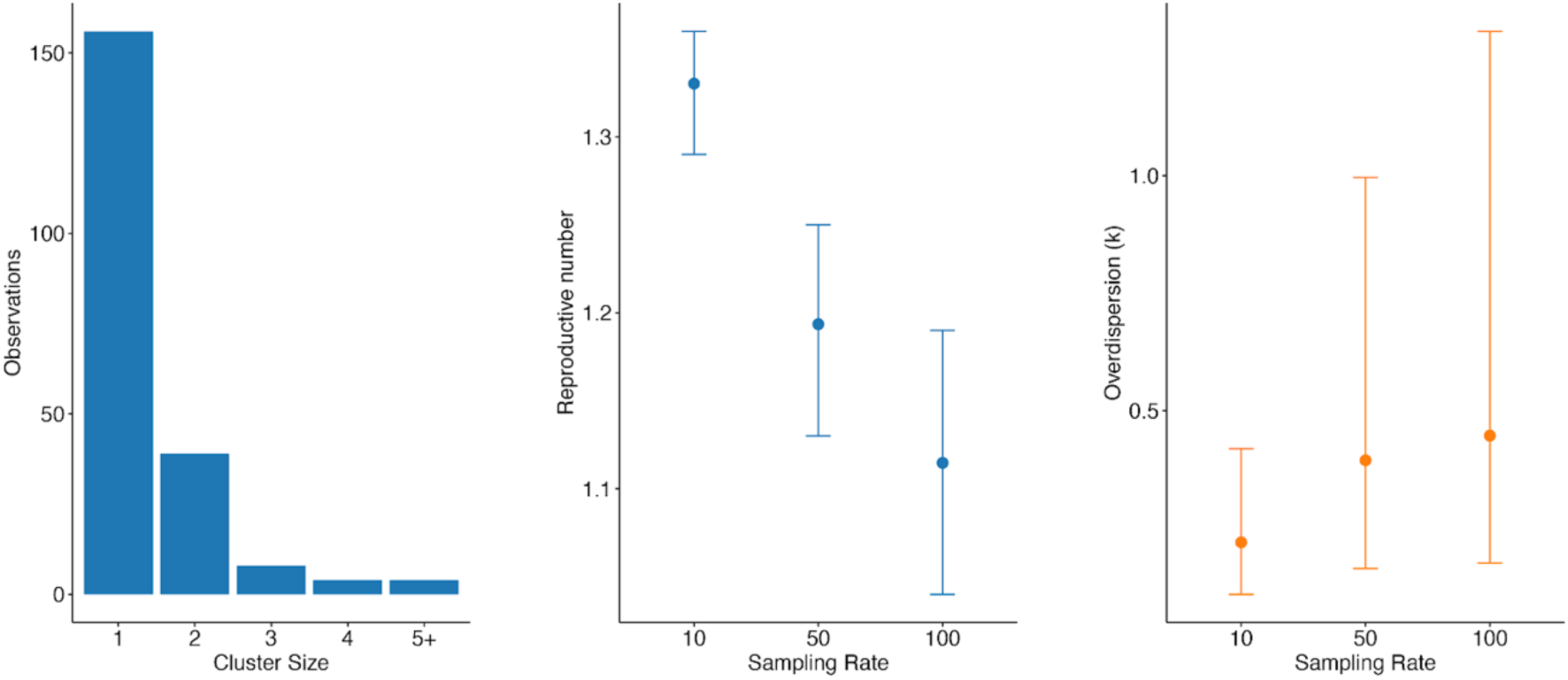
Distribution of the size of clusters of identical sequences during the mpox outbreak. Estimates for the reproduction number R and the dispersion parameter k assuming that 10, 50 and 100% were detected as cases.

## Notes

### Competing Interest Statement

The authors have declared no competing interest.

### Author Declarations

This study was conducted as part of the public health response to the mpox outbreak under the mandate of the National Public Health Agency (NPHA), Ministry of Health, Sierra Leone. All activities were carried out in accordance with national outbreak investigation guidelines. As this investigation formed part of an emergency public health response, it was exempt from institutional ethics review and the requirement for individual informed consent under national public health regulations. All data were anonymized prior to analysis to protect patient confidentiality.

